# Unveiling Alcohol Bias and Impaired Inhibitory Control in Young Binge Drinkers: Insights from the Alcohol Hayling Task

**DOI:** 10.1101/2025.01.10.25320346

**Authors:** Carina Carbia, Natália Almeida-Antunes, Margarida Vasconcelos, Eduardo López-Caneda

**Affiliations:** Louvain Experimental Psychopathology Research Group (UCLEP), Psychological Sciences Research Institute (IPSY), UCLouvain, Belgium; Center for Translational Health and Medical Biotechnology Research (TBIO), Health Research Network (Rise-Health), Polytechnic University of Porto, Portugal; Psychological Neuroscience Laboratory, Psychology Research Center (CIPsi), School of Psychology, University of Minho, Portugal

**Keywords:** alcohol, binge drinking, alcohol bias, craving, inhibitory control, semantic network

## Abstract

Craving and alcohol-related bias play a central role in addiction development. Previous research suggests that individuals with alcohol misuse exhibit heightened alcohol bias and deficits in inhibitory control, contributing to increased craving and sustained alcohol consumption. However, this relationship remains poorly understood in young binge drinkers, particularly regarding a specific form of alcohol bias known as semantic alcohol bias (heightened automatic accessibility of alcohol-related concepts), which may influence craving and drinking behavior. The present study is aimed at addressing this gap. A total of 81 college students (41 Binge Drinkers and 40 Non/Low Drinkers). completed craving questionnaires and the Alcohol Hayling task, a sentence-completion paradigm designed to measure semantic alcohol bias and inhibitory control. Results revealed that young binge drinkers generated more alcohol-related words in drinking- context sentences, committed more errors (i.e., producing alcohol-related words when they should have generated unrelated words), and displayed slower response times when inhibiting alcohol-related responses. A positive correlation was observed between craving and the frequency of alcohol-related words. These findings suggest that young binge drinkers exhibit a semantic accessibility bias towards alcohol-related concepts and difficulties inhibiting alcohol-related content. This study highlights the role of alcohol-related semantic networks in craving states, providing new insights into how alcohol biases may contribute to binge drinking behaviors among youth.

## Introduction

Binge drinking (BD) is characterized by a pattern of heavy alcohol consumption interspersed with periods of abstinence, a behavior highly prevalent among adolescents and young adults in most Western countries [1]. BD is typically defined as the intake of five or more alcoholic beverages within approximately two hours for men and four or more beverages for women [2, 3]. These recurring episodes of intoxication have been linked to various neurocognitive impairments [4, 5], including difficulties in inhibitory control and heightened alcohol bias.

Inhibitory control refers to the cognitive process involved in suppressing irrelevant information or overriding automatic responses that interfere with the execution of an ongoing task [6, 7]. Alcohol bias, on the other hand, is defined as the preferential automatic processing of alcohol-related cues (e.g., enhanced attention allocation, faster approach tendencies), reflecting their heightened incentive motivational properties [8, 9]. Both reduced inhibitory control and alcohol bias are linked to an increased risk of developing alcohol use disorder (AUD) [10, 11] and play a critical role in understanding addictive behaviors [12]. Accordingly, several neurobiological models of addiction — such as the triadic neurocognitive approach, the somatic marker theory, and the impaired Response Inhibition and Salience Attribution (iRISA) model —suggest that addictive behaviors arise from a dysregulation of the brain’s reward and control systems, leading to heightened salience of drug-related cues and impaired response inhibition [13–15]. Specifically, alcohol bias illustrates the aberrant salience attribution observed in alcohol-dependent individuals and heavy drinkers, where alcohol-related stimuli gain prominence and compelling motivational properties [16–18]. Inhibitory control, in turn, plays a crucial role in the emergence of biases, as response inhibition deficits reduce the capacity to suppress automatic, prepotent responses to alcohol-related cues, which could lead to difficulties in resisting drinking urges [19–21].

Another key factor contributing to the risk of alcohol misuse is craving, which is defined as the intense urge or desire to consume alcohol. Craving can perpetuate ongoing alcohol consumption [9, 22] and has been closely associated with alterations in inhibitory control and alcohol bias. Craving not only promotes preferential processing of alcohol-related stimuli [23], but also reinforces cognitive disruptions, further diminishing the ability to resist alcohol-related cues and exert self-control [24, 25]. The significance of craving in relation to alcohol attentional bias is underscored by findings in nonclinical populations, such as binge or social drinkers, where the intensity of craving often predicts alcohol bias more accurately than the quantity of alcohol consumed [10, 17, 26].

One type of alcohol bias that has been scarcely investigated is the semantic accessibility bias [27, 28], which —for the specific contex of alcohol use- involves enhanced automatic mental access to alcohol-related language [29]. Through repeated alcohol-related experiences, the maladaptive motivational memories —memories associated with alcohol use that reinforce harmful behavioral patterns- typical from heavy alcohol users [30–32], may strengthen the properties of the alcohol-related semantic network, leading to greater automatic accessibility of alcohol-related thoughts [33]. In this context, semantic accessibility renders alcohol-related cues and thoughts more mentally dominant, likely impacting inhibitory control and increasing the urge to consume alcohol or experience craving, which may contribute to the perpetuation and/or escalation of drinking patterns [34–36]. A semantic accessibility bias, favoring the activation of the alcohol-related semantic network, has been reported in individuals with alcohol misuse [29]; however, no studies have yet investigated this phenomenon in young BDs.

Paradigms designed to assess the individual’s semantic networks –rather than relying on predefined cues- could be particularly useful for studying alcohol bias and craving in the young binge drinking population. For example, craving might be triggered by specific semantic networks in patients with alcohol ude disorder compared to young binge drinkers (e.g., popular alcoholic beverages among young people or associations with activities like partying and celebrations). Therefore, in this study, we will employ a sentence-completion paradigm [6] to investigate alcohol-related semantic networks, specifically using the Alcohol Hayling Task [29]. Through highly constrained sentence frames and emphasis on speeded responses, this task elicits an automatic (drinking-related) prepotent responses, having to complete a missing word strongly cued by the preceding context [37, 38]. In the Inhibition section, participants are required to suppress the prepotent response (i.e., a semantically appropriate word) and instead provide an unrelated, nonsensical response, requiring the inhibition of automatically activated drinking-related semantic associations [38, 39].

Thus, the primary aim of this study is to examine inhibitory control and alcohol bias in young binge drinkers, focusing on their alcohol-related semantic networks. We hypothesize that binge drinkers will exhibit an alcohol-related semantic bias (e.g., increased production of alcohol-related words) and impaired inhibitory control (e.g., slower response times when inhibiting alcohol-related responses) compared to non-drinkers or low drinkers. Additionally, we anticipate that semantic alcohol bias will correlate with increased craving levels and/or alcohol use, aligning with the notion that semantic networks may contribute to maintaining BD behavior.

## Material and Methods

### Participants

Participants were contacted via email sent through the university’s email service, and a convenience sample of college students at the University of Minho (Braga, Portugal) was recruited. They completed a survey assessing sociodemographic variables (e.g., age, sex) and psychological characteristics using instruments such as the Barratt Impulsiveness Scale (BIS-11) and the Symptoms Checklist-90-Revised (SCL-90-R). A comprehensive evaluation of alcohol consumption habits was conducted, including the average number of drinks consumed in a standard week, the maximum number of drinks consumed on a single occasion in a typical week, the drunkenness ratio (i.e., the percentage of drinking episodes resulting in drunkenness, defined as loss of motor/verbal coordination, loss of self-control, and/or nausea), consumption speed (measured in units per hour), frequency of alcohol consumption (never, less than once per month, at least once per month, at least once per week), and age of onset of alcohol use and the first episode of BD. Participants also completed the Alcohol Use Disorder Identification Test (AUDIT) [40], a globally recognized self-report measure designed to identify current harmful and hazardous drinking patterns. Additionally, participants completed the Alcohol Craving Questionnaire Short-Form Revised (ACQ-SF-R) [41] and the Penn Alcohol Craving Scale (PACS), aimed at assessing short- and medium-term alcohol craving, respectively [42, 43].

Exclusion criteria for both groups included the presence of: family history of substance abuse (including alcoholism); use of illegal drugs except cannabis (i.e., at most once a month); alcohol abuse (i.e., score ≥ 20 in the AUDIT); personal history of psychopathological disorders (according to DSM-5 criteria); consumption of medical drugs with psychoactive effects (e.g., sedatives or anxiolytics) in the two weeks prior to the experiment; history of traumatic brain injury or neurological disorders; occurrence of an episode of loss of consciousness lasting > 20 min; score ≥ 90 in the Global Severity Index of the Symptom Checklist-90-Revised questionnaire [44] or in at least two of its symptomatic dimensions; and non-corrected sensory deficits. Two groups of participants were created: non/low drinkers and binge drinkers (see Table 1). Students who reported (i) drinking 5 or more drinks on one occasion at least once a month and (ii) drinking at a rate of at least two drinks per hour during these episodes were classified as binge drinkers. Those who reported (i) never drinking 5 or more drinks on each occasion and (ii) an AUDIT score ≤ 4 were considered non/low drinkers. Eighty-one participants (75 right-handed) were selected to take part in the experiment: 41 binge drinkers and 40 non/low drinkers. The study received approval by the Institutional Ethics Committee for Social Sciences and Humanities of the University of Minho, Braga, Portugal (CECSH 078/2018).

**Table 1.**
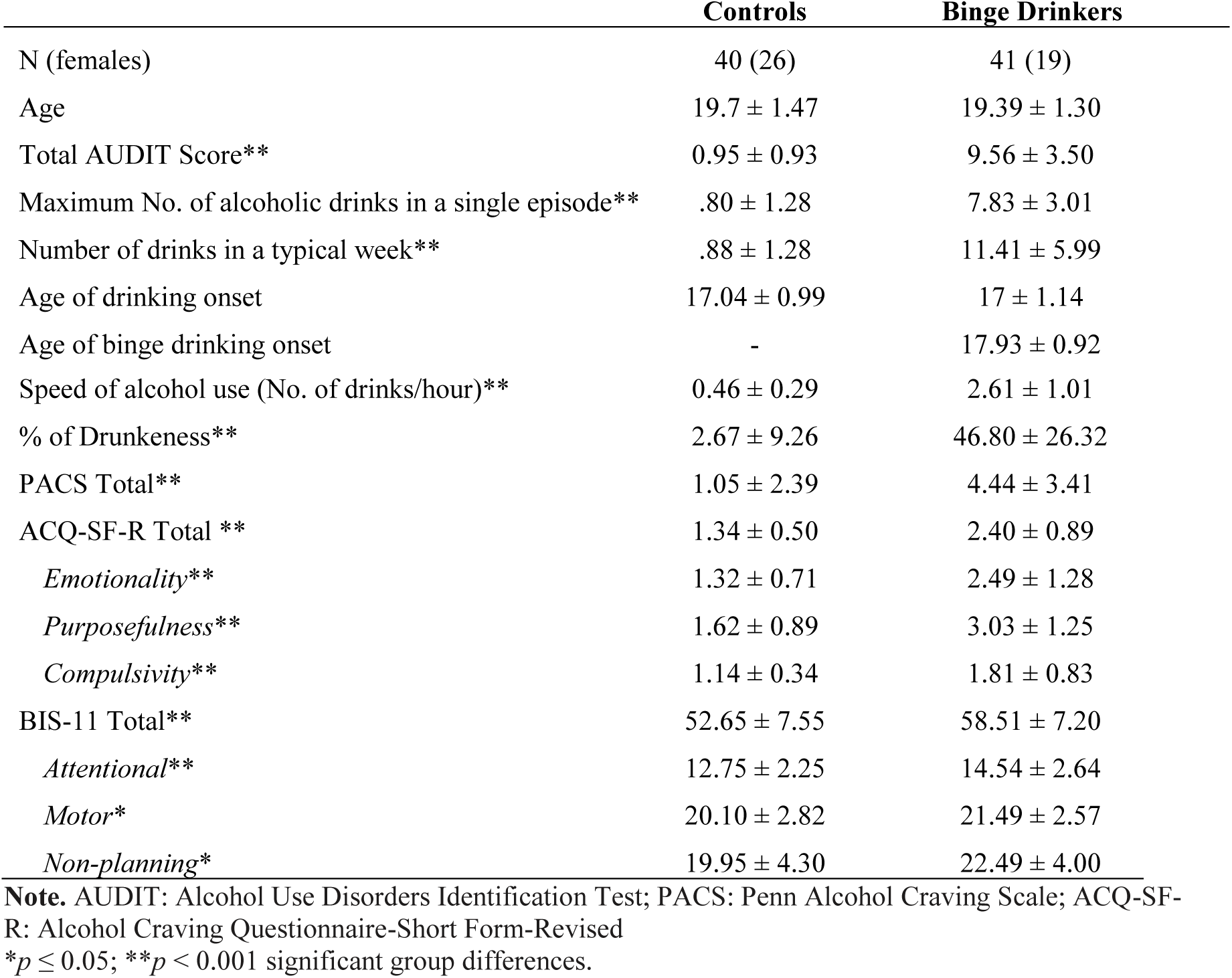
Sociodemographic and drinking characteristics of the sample (mean ± SD).

## Procedure

### Alcohol Hayling task

Initially designed by Burgess and Shallice (1997) to assess executive dysfunction in patients, the adapted Alcohol Hayling task is a sentence-completion task derived from the work of Rose et al. [29] that measures alcohol-related bias and inhibitory control. This paradigm consists of sentences in which the final words are omitted, but there is a particularly high likelihood of one specific response. The task, designed by Rose and cols. and adapted to Portuguese by our research team, comprises two different conditions (Neutral and Drinking) that were counterbalanced across participants. Each of these conditions has two sequential phases: (a) Initiation and (b) Inhibition. At the Initiation phase (section A), which assesses general automatic responses, participants were read 15 incomplete sentences and asked to complete each of them as quickly as possible, with the first word which comes to mind. The word is strongly cued by the preceding context, and the individual has to complete the sentence by producing the missing word (e.g., in the sentence “This man has travelled everywhere around the…[*world*]”) [37]. Subsequently, during the Inhibition phase (section B), participants were read the same sentences and asked to complete them with an unrelated word or a word that makes no sense in that context (i.e., inhibit a semantically correct word); for example, “This man has travelled everywhere around the…[*shark*]”.

On the other hand, during the Initiation phase of the Drinking condition, similar to the Neutral condition, participants were required to complete the sentences with the first word that came to mind that made semantic sense. However, unlike the Neutral condition, all the sentences were set within a context of beverage consumption, so the generated words had to be related to drinking or beverages, albeit there was no direct priming for alcohol-related responses (e.g., “She drank a glass of… [*wine/water*]”). In the Inhibition phase, participants were instructed to provide a word that did not make sense at all (e.g., “She drank a glass of… [*broom*]”. In this phase, for both the Neutral and Drinking conditions, responses were scored as *error type 1* if the word made sense within the sentence, *error type 2* if it did not make sense but was semantically related to the sentence, and *correct response* if it made no sense at all.

In the Initiation phase, participant’s responses informed about an accessibility bias towards alcohol-related semantic networks in sentence completion, indicating the activation of alcohol-related cognitions. In the Inhibition phase, the task measures the ability to suppress automatic or pre-potent (alcohol- and nonalcohol-related) responses. By comparing the Neutral and Drinking conditions, the task helps to isolate inhibitory control deficits specifically associated with alcohol-related content.

The mean reaction times to word generation, the inhibition latency (i.e., the subtraction of the total time taken to respond in section B and the total time taken to respond in section A) and the total number of errors and correct responses in section B were estimated for both the Neutral and Drinking conditions. In addition, the number of alcohol-related words given across all sentences in the Drinking condition was also calculated.

### Statistical analyses

The statistical analyses were performed using IBM SPSS Statistics for Macintosh v28.0.0.0 (IBM Corp., Armonk, NY). Between-group comparisons (i.e., independent samples *t*-tests) were calculated to examine potential group differences regarding drinking (e.g., AUDIT scores, number of drinks consumed in a typical week, age of onset of alcohol use) and psychological (e.g., self-reported impulsivity and craving levels) characteristics. Additionally, the same between-group analysis was conducted to explore task performance, including reaction times, inhibition latency, correct and incorrect responses, and the number of alcoholic words. Correlational analyses were also used to test potential associations between Hayling task performance and drinking and craving measures.

## Results

### Substance use and psychological variables

There were significant between-group differences in the following substance use variables: Total AUDIT score [t(79) = -15.01, p < .001], maximum number of alcoholic drinks in a single episode [t(79) = -16.01, p < .001], number of drinks in a typical week [t(79) = -10.88, p < .001], speed of alcohol use [t(79) = -14.57, p < .001], and percentage of drunkenness [t(79) = -10.00, p < .001]. In addition, groups also differed with regard to craving measures: PACS total score [t(79) = -5.16, p < .001], ACQ total score [t(79) = - 6.58, p < .001], ACQ Emotionality [t(79) = -5.03, p < .001], ACQ Purposefulness [t(79) = -5.82, p < .001], ACQ Compulsivity [t(79) = -4.73, p < .001]. In all of them, binge drinkers showed increased values in comparison to the control group (see Table 1).

### Behavioral results

While there were no significant between-group differences regarding the Neutral condition, results revealed that binge drinkers generated more alcohol-related words during the Initiation phase (part A) of the Drinking condition [t(79) = -2.22, p = .014] (see Figure 1), and they made more type 2 incorrect responses (i.e., they provide more semantically alcohol-related words when they should be generating words that are not semantically related to the phrase; e.g.: "She asked if he wanted a can of …", Answer: *drink*) [t(79) = -1.78, p = .039] (see Figure 2). In addition, binge drinkers exhibited longer inhibition latency than their control peers [t(79) = -1.82, p = .036] (see Figure 3), meaning they took more time to suppress the production of alcohol-related words relative to neutral words [t(79) = -1.82, p = .036].

**Figure 1.**
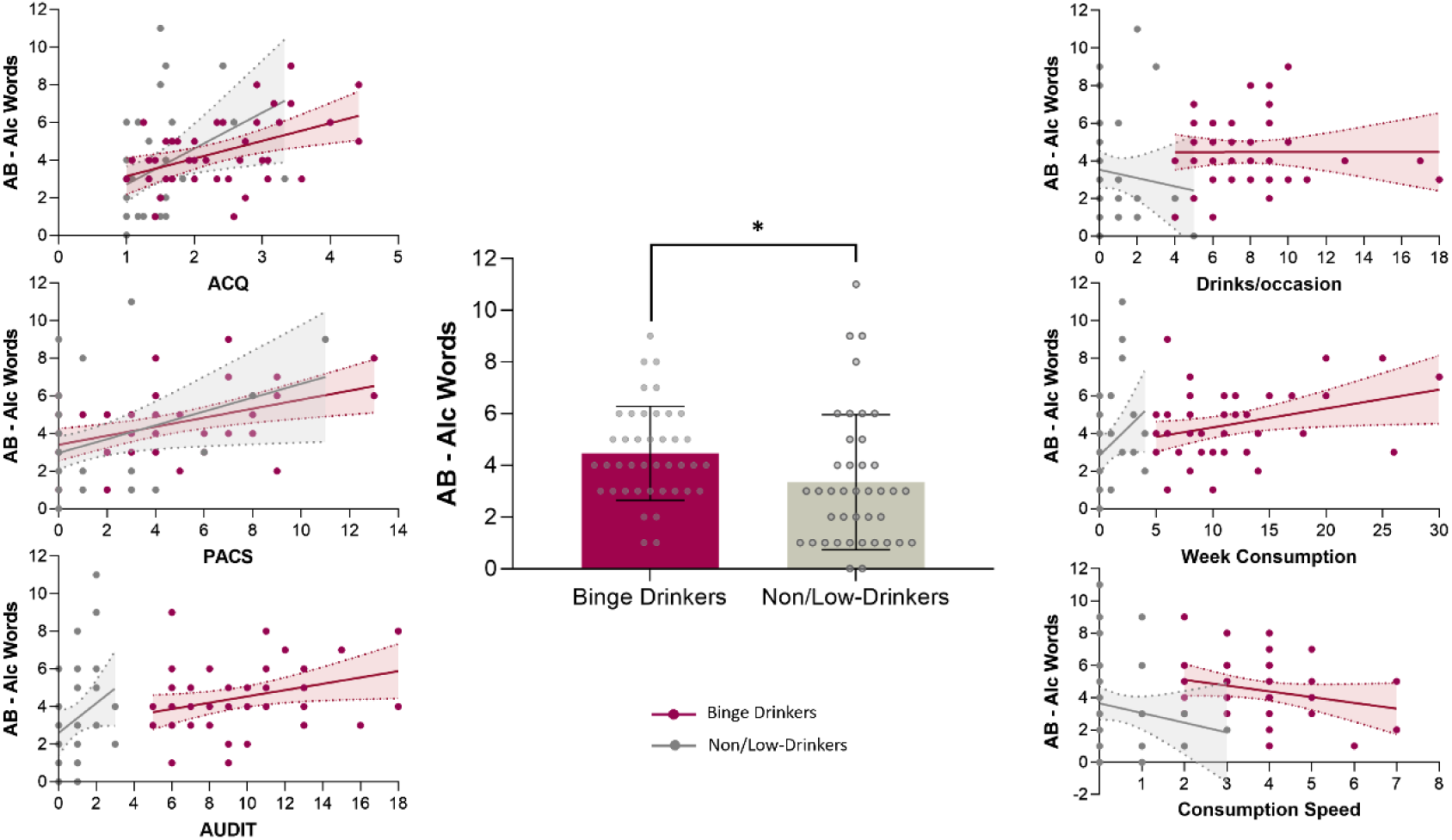
Central bar graph illustrating group differences in the number of alcohol-related words produced during the drinking condition, comparing Non/Low-Drinkers (gray) and Binge Drinkers (bordeaux). Scatter plots on the left and right show the distribution of this measure in relation to craving indices (PACS and ACQ) and drinking behaviors.

**Figure 2.**
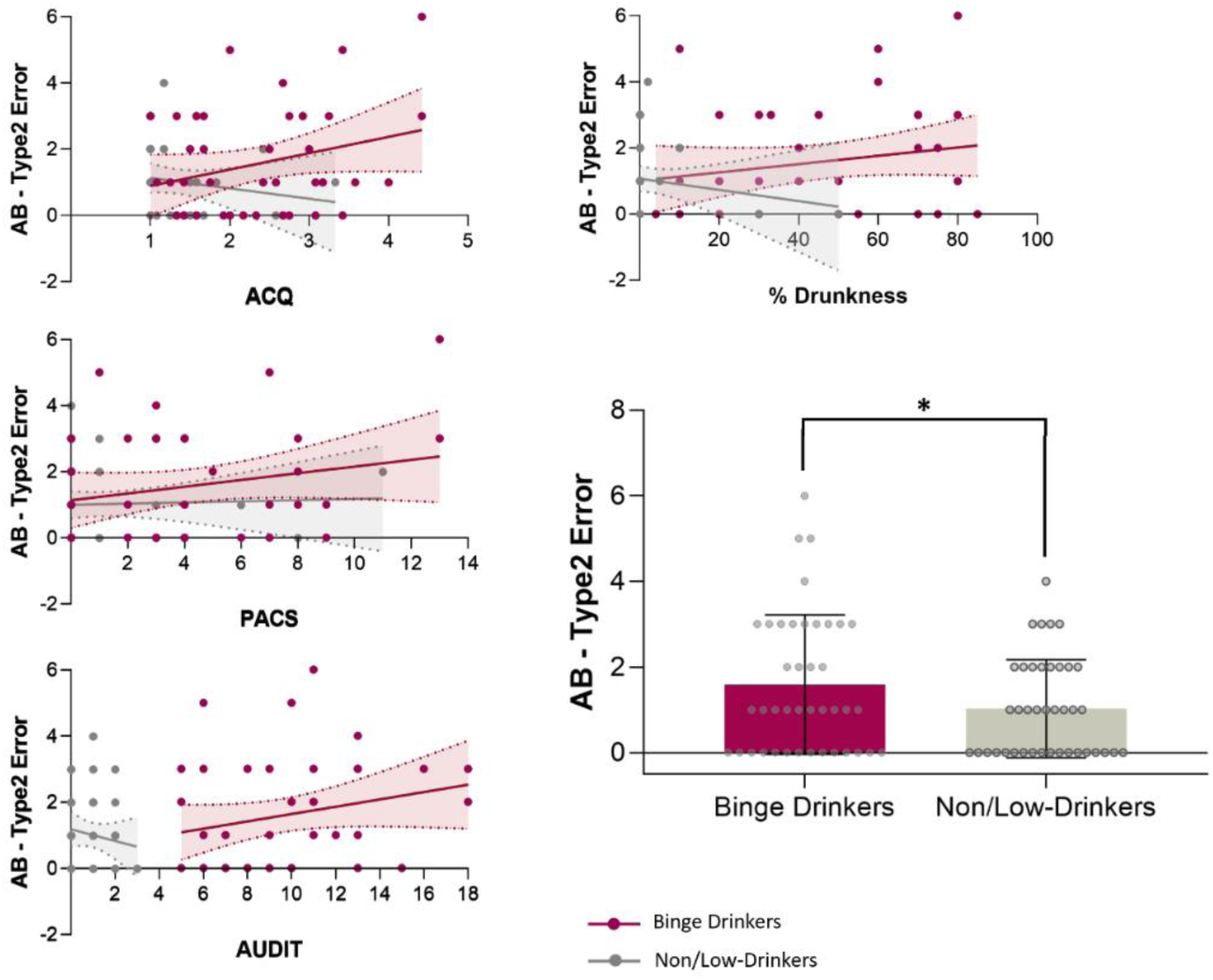
Bar graph (right) illustrating group differences in the number of type 2 incorrect responses between Non/Low-Drinkers (gray) and Binge Drinkers (bordeaux). Scatter plots (left) display the distribution of this measure in relation to craving indices (PACS and ACQ) and the total AUDIT score.

**Figure 3.**
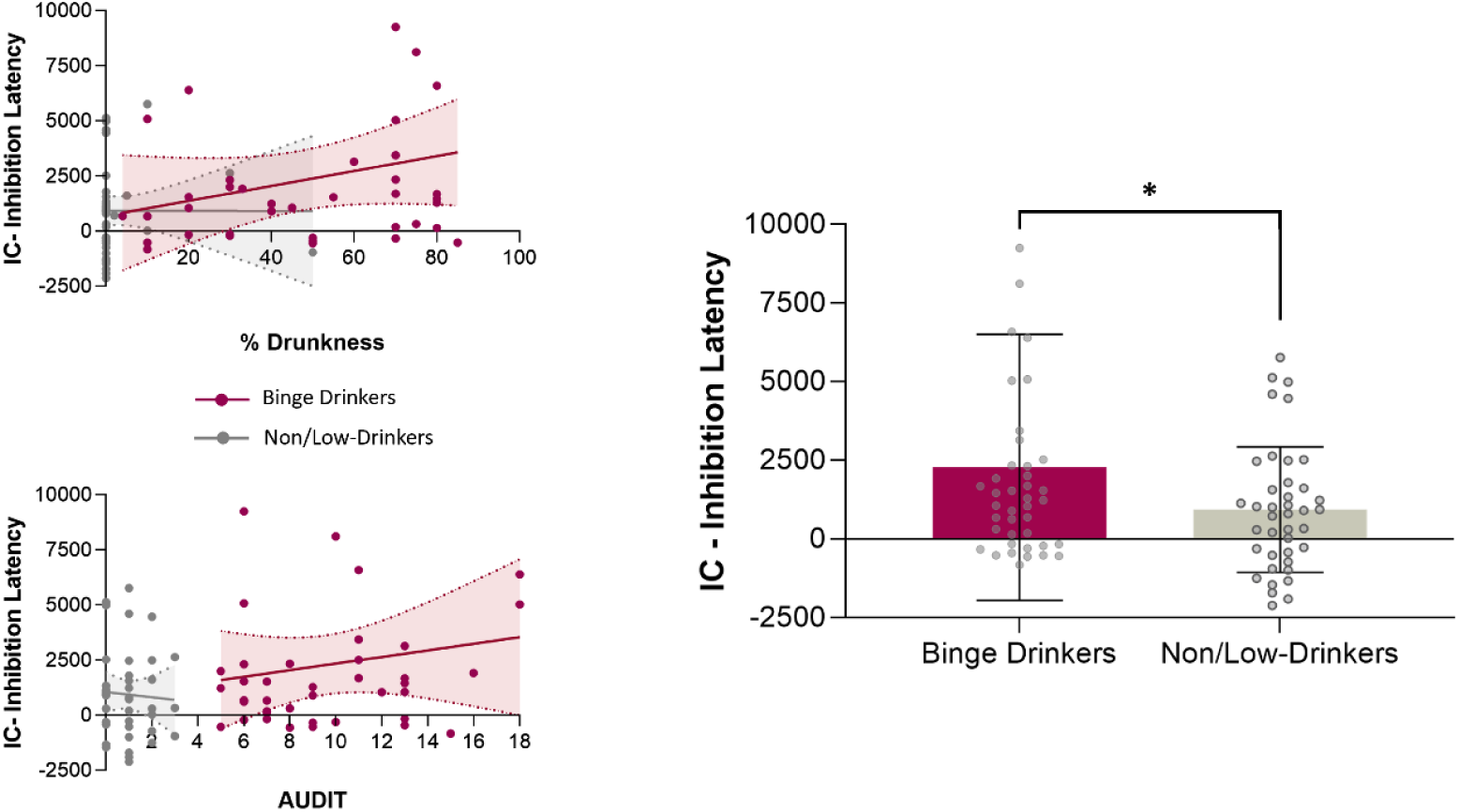
Bar graph (right) illustrating group differences in inhibition latency between Non/Low-Drinkers (gray) and Binge Drinkers (bordeaux). Scatter plots (left) show the distribution of this measure in relation to the percentage of drunkenness and the total AUDIT score.

### Correlational analysis

Correlation analyses were performed between drinking and craving variables and the behavioral measures that showed group differences in the main analysis (see Figure 4). There were significant positive associations between the number of alcoholic words produced during the drinking condition and the AUDIT total score (r = .327, p = .003), the number of drinks in a typical week (r = .333, p = .002), the maximum number of alcoholic drinks in a single episode (r = .233, p = .036), the speed of alcohol drinking rate (r = .242, p = .030), the PACS total score (r = .432, p < .001), and the ACQ total score (r = .437, p < .001). The number of type 2 incorrect responses was positively correlated with the AUDIT total score (r = .254, p = .022), the percentage of drunkness (r = .235, p =.036), the PACS total score (r = .232, p = .038), and the ACQ total score (r = .239, p = .032). Finally, the inhibition latency was positively associated with the AUDIT total score (r = .224, p = .044), and the percentage of drunkness (r = .264, p = .018).

**Figure 4.**
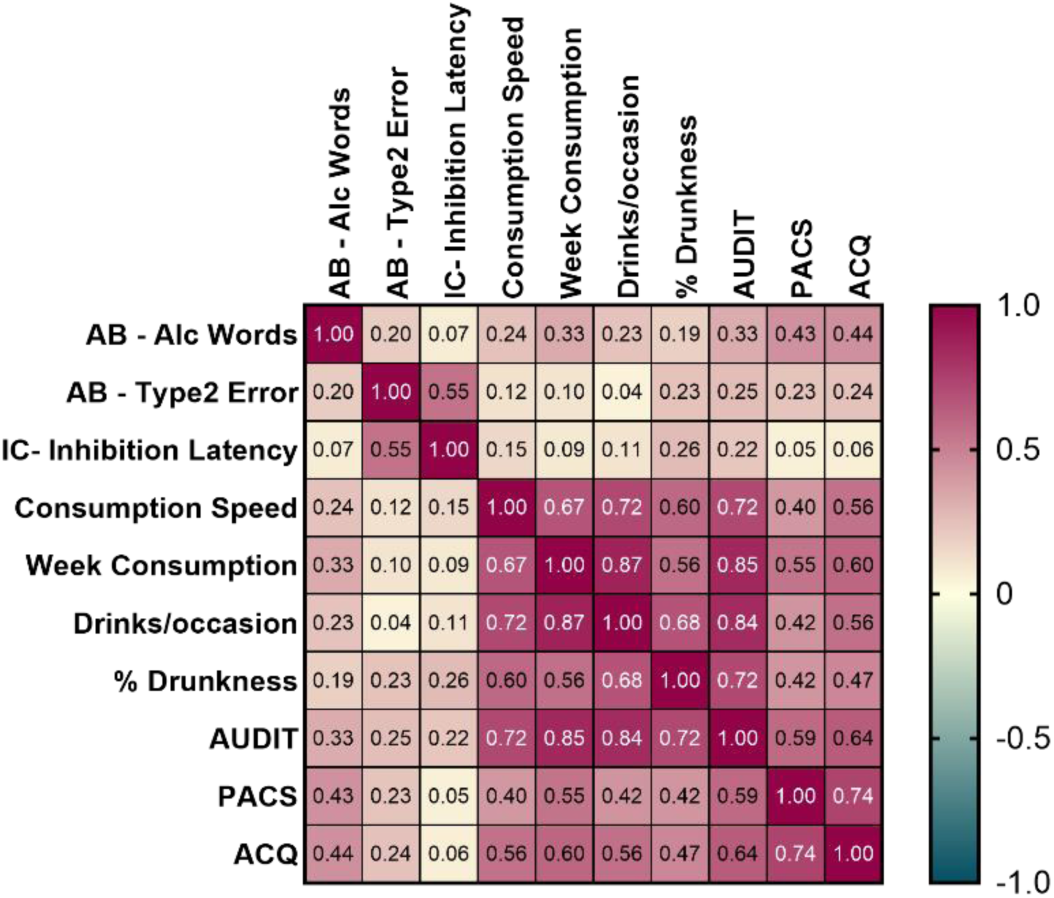
Correlation matrix between Alcohol Bias (number of alcoholic words and type 2 erros), Inhibitory Control (inhibition latency), and craving and alcohol consumption measures. Note: AB – Alcohol Bias; IC – Inhibitory Control.

## Discussion

The present study was the first to explore alcohol accessibility bias and the inhibition of alcohol-related semantic networks in youths with a BD pattern. The findings revealed that, compared to their control peers, young binge drinkers generated more alcohol-related words during the initiation phase in Drinking contexts, committed more *Type 2 errors* (e.g., producing alcohol-related words when the task required nonsensical responses), and displayed slower reaction times when inhibiting drinking-related words. Notably, no difficulties were observed in initiation, suppression, or strategy generation in Neutral contexts. These results suggest an alcohol-specific semantic bias, coupled with difficulties in inhibiting prepotent alcohol-related responses.

The present findings align with previous studies in individuals with harmful alcohol use patterns demonstrating inhibitory deficits using the original Hayling task. Accordingly, patients with AUD showed slower reaction times for the inhibitory part and increased inhibition errors compared to controls [34, 45, 46]. These inhibitory deficits are consistent with studies showing poorer performance and/or neurofunctional anomalies in young binge drinkers –relative to non- or low-drinking peers- across inhibitory control paradigms such as the Stroop, Stop Signal, and Go/No-Go tasks [47–51].

Our results support the notion of inhibitory deficits in young individuals with a BD pattern, extending previous findings by revealing, for the first time, that these abnormalities also manifest at the level of alcohol-related semantic networks. This impairment in inhibiting alcohol-related semantic associations may reinforce alcohol-related thoughts and, ultimately, contribute to sustained alcohol use in this population. Notably, in line with this hypothesis, a recent study found that binge drinkers exhibit abnormal brain functional connectivity patterns –specifically, hyperconnectivity between inhibitory control and memory networks- when suppressing alcohol-related memories, suggesting heightened attention to intrusive alcohol-related cues and increased engagement of top-down control mechanisms to suppress these intrusions [30]. Similarly, the longer response latencies observed in the BD group when inhibiting alcohol-related words in ambiguous (i.e., alcohol or non-alcohol) drinking contexts may indicate poor inhibitory control and/or greater cognitive effort required to suppress alcohol-related semantic associations [28, 29, 37]. Such difficulties in suppressing the activation of alcohol-related semantic networks may, in turn, reinforce a cycle where thoughts or memories of alcohol are more easily activated, potentially sustaining drinking behaviors. Alongside the observed inhibitory control difficulties, our results indicate the presence of semantic alcohol bias in young binge drinkers, as they were more likely to complete sentences with alcohol-related words during the Initiation phase. For example, in ambiguous drinking-context sentences, such as “The man bought a liter of…”, binge drinkers frequently provided alcohol-related completions (e.g., “beer,” “wine”), whereas controls tended to use neutral terms (e.g., “water,” “milk”). Given that sentences must be completed as quickly as possible, minimizing controlled processing and encouraging automatic responses, this finding suggests a greater automatic accessibility for alcohol-related semantic associations, likely shaped by prior experiences with alcohol and heightened craving levels [28, 29]. Supporting this, a greater number of alcohol-related words positively correlated with craving scores as well as with key features of BD behavior such as maximum number of alcoholic drinks in a single episode and drinking speed.

Similar patterns have been observed in non-clinical alcohol misuse populations employing other alcohol bias paradigms, such as the Alcohol Stroop task [11, 53], Flanker task [54], and Visual Probe task [18]. Additionally, techniques such as eye-tracking [10, 55] electroencephalography [56], and functional magnetic resonance imaging [57] have also demonstrated a bias towards alcohol stimuli in individuals with alcohol misuse. Importantly, our findings also align with those of Rose and colleagues [29], the only other study using the Alcohol Hayling Task. In their work, heavy-drinking adults exhibited a marked tendency to generate alcohol-related responses and longer response latencies during inhibition, reflecting alcohol-related inhibitory difficulties and a semantic accessibility bias [29].

Collectively, our results and previous evidence suggest that strong alcohol-related associations, driven by the heightened salience of alcohol cues [58], may become readily accessible within the semantic network, particularly in ambiguous or socially permissive drinking contexts such as parties, gatherings with friends or celebrations [33]. Repeated experiences of heavy alcohol consumption may further strengthen maladaptive motivational memories, reinforcing the link between alcohol cues and substance-seeking behaviors through persistent neuroadaptive changes in the brain’s reward system [31, 59]. This increased accessibility likely increases the frequency of alcohol-related thoughts, which, combined with inhibitory deficits, may perpetuate or escalate drinking behaviors [60]. In particular, in individuals with alcohol use disorder, enhanced memory for alcohol-related associations has been shown to predict heavier real-world drinking [61] and stronger cue-elicited cravings [62]. Even among subclinical populations, such as binge drinkers, the heightened salience of alcohol-related associations may increase their accessibility within the semantic network, reinforcing a cycle in which easily accessible alcohol-related thoughts, combined with inhibitory deficits, intensify cravings. Nonetheless, disentangling the causal interplay between craving and semantic bias requires further investigation to better understand their underlying mechanisms.

In conclusion, the current study revealed that young binge drinkers exhibit heightened accessibility to alcohol-related semantic networks alongside notable difficulties in inhibiting these associations, which may help sustain craving and BD behaviors. Additionally, these findings suggest semantic networks might be a promising focus for cognitive bias modification [63] and/or cognitive training strategies [64] in this population.

## Ethics

The study received approval by the Institutional Ethics Committee for Social Sciences and Humanities of the University of Minho, Braga, Portugal (approval reference: CECSH 078/2018). All aspects of research were followed in accordance with the Declaration of Helsinki.

## Data Availability

All data produced in the present study are available upon reasonable request to the authors

## Acknowledgements

The study was conducted at the Psychology Research Center (PSI/ 01662), School of Psychology, University of Minho, supported by the Foundation for Science and Technology (FCT) through the Portuguese State Budget (Ref.: UIDB/PSI/01662/2020). Eduardo López-Caneda received support from the FCT and the Portuguese Ministry of Science, Technology and Higher Education, through the Individual Call to Scientific Employment Stimulus (CEECIND/07751/2022), and by the project PTDC/PSI-ESP/ 1243/2021. Carina Carbia received support from the Fund for Scientific Research *(Charge de Recherche, Fonds de la Recherche Scientifique [FNRS])*.

We confirm that neither the authors have received any payments or services from a third party in the past 36 months, or at any time, that could be perceived to influence the submitted work.

## Conflict of Interest

The authors declare no conflict of interest.

## Notes

### Competing Interest Statement

The authors have declared no competing interest.

### Author Declarations

The study received approval by the Institutional Ethics Committee for Social Sciences and Humanities of the University of Minho, Braga, Portugal (approval reference: CECSH 078/2018).

